# Distinguish Coronavirus Disease 2019 Patients in General Surgery Emergency by CIAAD Scale: Development and Validation of a Prediction Model Based on 822 Cases in China

**DOI:** 10.1101/2020.04.18.20071019

**Authors:** Bangbo Zhao, Yingxin Wei, Wenwu Sun, Cheng Qin, Xingtong Zhou, Zihao Wang, Tianhao Li, Hongtao Cao, Weibin Wang, Yujun Wang

**Affiliations:** Department of General Surgery, Peking Union Medical College Hospital, Peking Union Medical College, Chinese Academy of Medical Sciences, Beijing, 100005, China; Department of Surgery, Peking Union Medical College Hospital, Peking Union Medical College, Chinese Academy of Medical Sciences, Beijing, 100005, China; Department of Critical Care Medicine, The Central Hospital of Wuhan, Tongji Medical College, Huazhong University of Sciences and Technology, Wuhan, Hubei Province, China

**Author notes:** Correspondence to: Weibin Wang, M.D., Ph.D. (Vice Director, Professor; Department of General Surgery, Peking Union Medical College Hospital, Shuaifuyuan 1, Doncheng District, Beijing, 100005, China; E-MAIL) and Yujun Wang, M.D., Ph.D. (Professor, Department of Critical Care Medicine, The Central Hospital of Wuhan, Tongji Medical College, Huazhong University of Science and Technology, Wuhan, Hubei Province, 430014, China; E-MAIL). ZBB, WYX, SWW and QC contributed equally to this article.

## Abstract

**IMPORTANCE:** In the epidemic, surgeons cannot distinguish infectious acute abdomen patients suspected COVID-19 quickly and effectively.

**OBJECTIVE:** To develop and validate a predication model, presented as nomogram and scale, to distinguish infectious acute abdomen patients suspected coronavirus disease 2019 (COVID-19).

**DESIGN:** Diagnostic model based on retrospective case series.

**SETTING:** Two hospitals in Wuhan and Beijing, China.

**PTRTICIPANTS:** 584 patients admitted to hospital with laboratory confirmed SARS-CoV-2 from 2 Jan 2020 to15 Feb 2020 and 238 infectious acute abdomen patients receiving emergency operation from 28 Feb 2019 to 3 Apr 2020.

**METHODS:** LASSO regression and multivariable logistic regression analysis were conducted to develop the prediction model in training cohort. The performance of the nomogram was evaluated by calibration curves, receiver operating characteristic (ROC) curves, decision curve analysis (DCA) and clinical impact curves in training and validation cohort. A simplified screening scale and managing algorithm was generated according to the nomogram.

**RESULTS:** Six potential COVID-19 prediction variables were selected and the variable abdominal pain was excluded for overmuch weight. The five potential predictors, including fever, chest computed tomography (CT), leukocytes (white blood cells, WBC), C-reactive protein (CRP) and procalcitonin (PCT), were all independent predictors in multivariable logistic regression analysis (p ≤0.001) and the nomogram, named COVID-19 Infectious Acute Abdomen Distinguishment (CIAAD) nomogram, was generated. The CIAAD nomogram showed good discrimination and calibration (C-index of 0.981 (95% CI, 0.963 to 0.999) and AUC of 0.970 (95% CI, 0.961 to 0.982)), which was validated in the validation cohort (C-index of 0.966 (95% CI, 0.960 to 0.972) and AUC of 0.966 (95% CI, 0.957 to 0.975)). Decision curve analysis revealed that the CIAAD nomogram was clinically useful. The nomogram was further simplified into the CIAAD scale.

**CONCLUSIONS:** We established an easy and effective screening model and scale for surgeons in emergency department to distinguish COVID-19 patients from infectious acute abdomen patients. The algorithm based on CIAAD scale will help surgeons manage infectious acute abdomen patients suspected COVID-19 more efficiently.

## Introduction

Since the outbreak of coronavirus disease 2019 (COVID-19) in Wuhan, China, which has been characterized as a pandemic by the World Health Organization on Mar 11, 2020, the cunning virus has drastically spread all over the world^1,2^. Millions of people have been infected, resulting in tens of thousands died^3^. The still ongoing pandemic is not only a huge threat to the public physical health but also an acid test for the medical system regardless of developed counties or developing countries^4^. In addition to prevention, quick and accurate diagnosis of COVID-19 is one of the most important tasks currently.

The medical management of other diseases has been critically disturbed, especially for some with fever, the typical symptom of COVID-19^5^. There are numerous high-risk people in close contact with the confirmed patients. As we all know, interdicting transmission is the most effective way to control the epidemic of COVID-19. Under current situation for surgeons, infectious acute abdomen is still one of the most common surgery emergencies, the patients of which often have fever, diarrhea and other atypical symptom, and also have similar change of blood routine and other biochemistry items with COVID-19 for infectious or chemical peritonitis^6^. Hence, when surgeons managing patients of infectious acute abdomen, the typical symptoms and blood test indicators of COVID-19 are easy to be covered up. Currently, diagnosis of COVID-19 mainly depends on severe acute respiratory syndrome coronavirus 2 (SARS-CoV-2) nucleicacid detection^7^. However, the approach has defects of time-consuming and false-negative results^8^, which go against the urgency of emergency operation and the aim to avoid cross infection in the hospital, and therefore, there is a pressing need of an easier and more feasible method to distinguish the genuine COVID-19 patients among the infectious acute abdomen patients with mimical symptoms.

Based on the clinical data of 822 patients, 584 of COVID-19 and 238 of infectious acute abdomen, we compared the demographic, clinical, imaging and laboratory characteristics to obtain significant predictors of COVID-19. Further, a prediction model to distinguish the two diseases was generated based on machine learning and presented in form of nomogram, which has a good discrimination performance in both training and validation cohort. Ultimately, we offered a practical screening scale, named CIAAD scale, and an algorithm, including precaution advice for surgeons, in infectious acute abdomen patients encounters.

## Methods

### Patients

Ethical approval was obtained from the Ethics Committees of Peking Union Medical College Hospital and the Central Hospital of Wuhan for this retrospective stud. We brought 584 COVID-19 patients adopted into the Central Hospital of Wuhan between Jan 2, 2020 and Feb 15, 2020 into our study, the diagnostic criteria of COVID-19 were positive RT-PCR results for SARS-CoV-2 or highly homologous viral gene sequencing results with SARS-CoV-2 of respiratory or blood samples^7^. Since the routine medical order of other diseases in Wuhan were severely disturbed by the epidemic, the clinical data of infectious acute abdomen patients were collected from 283 patients receiving emergency operation in Peking Union Medical College Hospital between Feb 28, 2019 and Apr 3, 2020. The inclusion criteria were: (1) fever; or (2) abnormal blood routine results or other infection indicators; or (3) signs of pneumonia. The patients with infectious acute abdomen adopted after Jan 20, 2020 were all tested for SARS-CoV-2 and none of them was positive.

### Data Collection and definitions

Demographic, clinical, laboratory, treatment and outcome data of the COVID-19 and infectious acute abdomen patients were extracted from electronic medical system of the Central Hospital of Wuhan and Peking Union Medical College Hospital respectively.

Fever was defined as axillary temperature of at least 37.3^°^C. The chest CT scores were graded retrospectively by two radiologists back to back. Each lung is divided into the upper, middle and lower parts and the scoring criterion is <1/3 lung infected, 0 point; 1/3-2/3 lung infected, 1 point; >2/3 lung infected, 2 points. The definition of COVID-19 severity was based on the Chinese management guideline for COVID-19 (version 7.0) published by the National Health Commission of China^7^.

### Potential predictors selection

The primary cohort of the whole 822 patients was divided into a training cohort and a validation cohort randomly by ratio of 2:1. The least absolute shrinkage and selection operator (LASSO) method, one of the most effective ones in regularized regression with great advantage when managing multicolinearity data, was used to select the most useful predictive variables for COVID-19 in the training cohort^9^.

### Development and validation of a prediction model

We conduct multivariate logistic regression involving the potential predictors to further verify their predictive efficacy and build a nomogram to distinguish COVID-19 patients from infectious acute abdomen patients on the basis of multivariable logistic analysis in the training cohort. We named the nomogram as COVID-19 and Infectious Acute Abdomen Distinguishment (CIAAD) nomogram. Calibration curves were plotted to assess the calibration of the CIAAD nomogram and C-index was measured to quantify its discrimination performance. Then, CIAAD nomogram the training cohort was applied to the patients in the validation cohort and the calibration curve and C-index were derived on the basis of the regression analysis. The distinguishment capacity of CIAAD nomogram in both training and validation cohorts were also accessed by calculating the area under the receiver operating characteristic curve (AUC).

### Clinical Usefulness Assessment

Decision curve analysis and clinical impact curves was conducted to evaluate the clinical practicability of CIAAD nomogram by quantifying the net benefits at different threshold probabilities in both training and validation dataset.

### Development of Screening Scale

The corresponding score of each item in CIAAD nomogram was divided by 25 and rounded to obtain a simplified score. The simplified score are verified to have the same efficacy comparing the original nomogram. We subdivide the risk of COVID-19 into low (<0.3), moderate (0.3-0.7) and high (>0.7) risk. The CIAAD Scale was formed on basis of the simplified scoring criterion and risk classification.

### Statistical analysis

Categorical variables were expressed as numbers and percentage. Continuous variables were expressed as medians with interquartile ranges. Chi-square test and Mann-Whitney U-test were used to evaluate categorical and continuous data respectively. Statistical analysis was conducted with R software (version 3.6.1; http://www.Rproject.org) and SPSS statistical software package (version 25.0). *P*<0.05 was considered statistically significant.

## Results

### Demographic and clinical characteristics

A total of 822 patients, 584 COVID-19 patients without infectious acute abdomen and 238 infectious acute abdomen patients without COVID-19, were included in this study (Table 1). Nearly 16% of the COVID-19 patients were severe or critical (Figure 1). The disease spectrum of the infectious acute abdomen patients was principally acute appendicitis (60.5%), perforation (17.6%) and obstruction (13.4%). The infectious acute abdomen patients are younger (P=.001) and combined with less chronic diseases, such as diabetes (P=.005), cardiovascular and cerebrovascular diseases (P=.011). Fever, as reported before, was the most common symptom in the COVID-19 patients (80.1%), which ranked second (32.4%) in the infectious acute abdomen patients inferior to abdominal pain (99.2%). COVID-19 resulted in larger infected occupancy of lung (P<.001). Nevertheless, abdominal infection caused by infectious acute abdomen rendered the laboratory testing results of these patients, such as CRP, PCT, WBC, neutrophils and fibrinogen, more abnormal (P<.001).

**Table 1.**
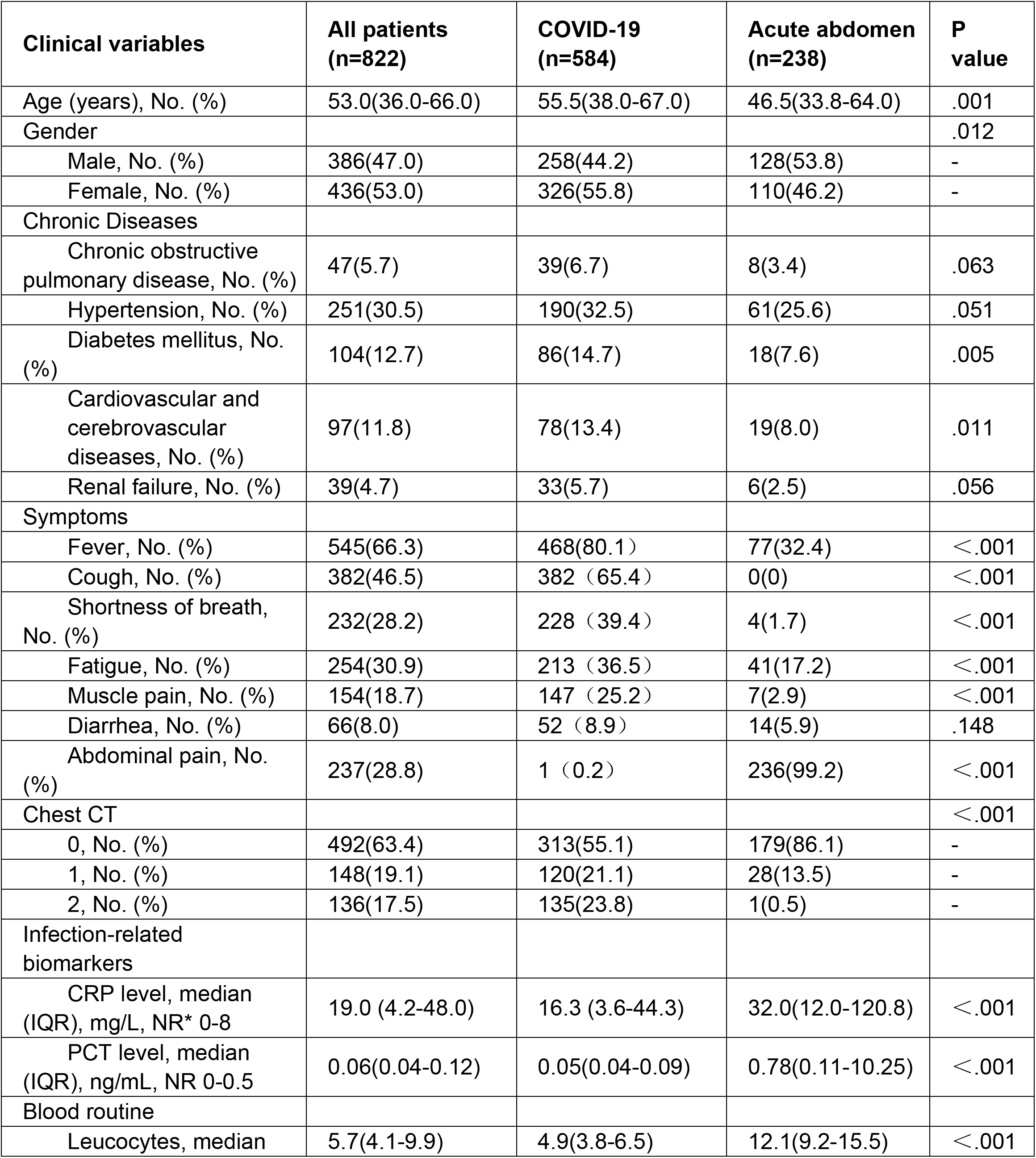
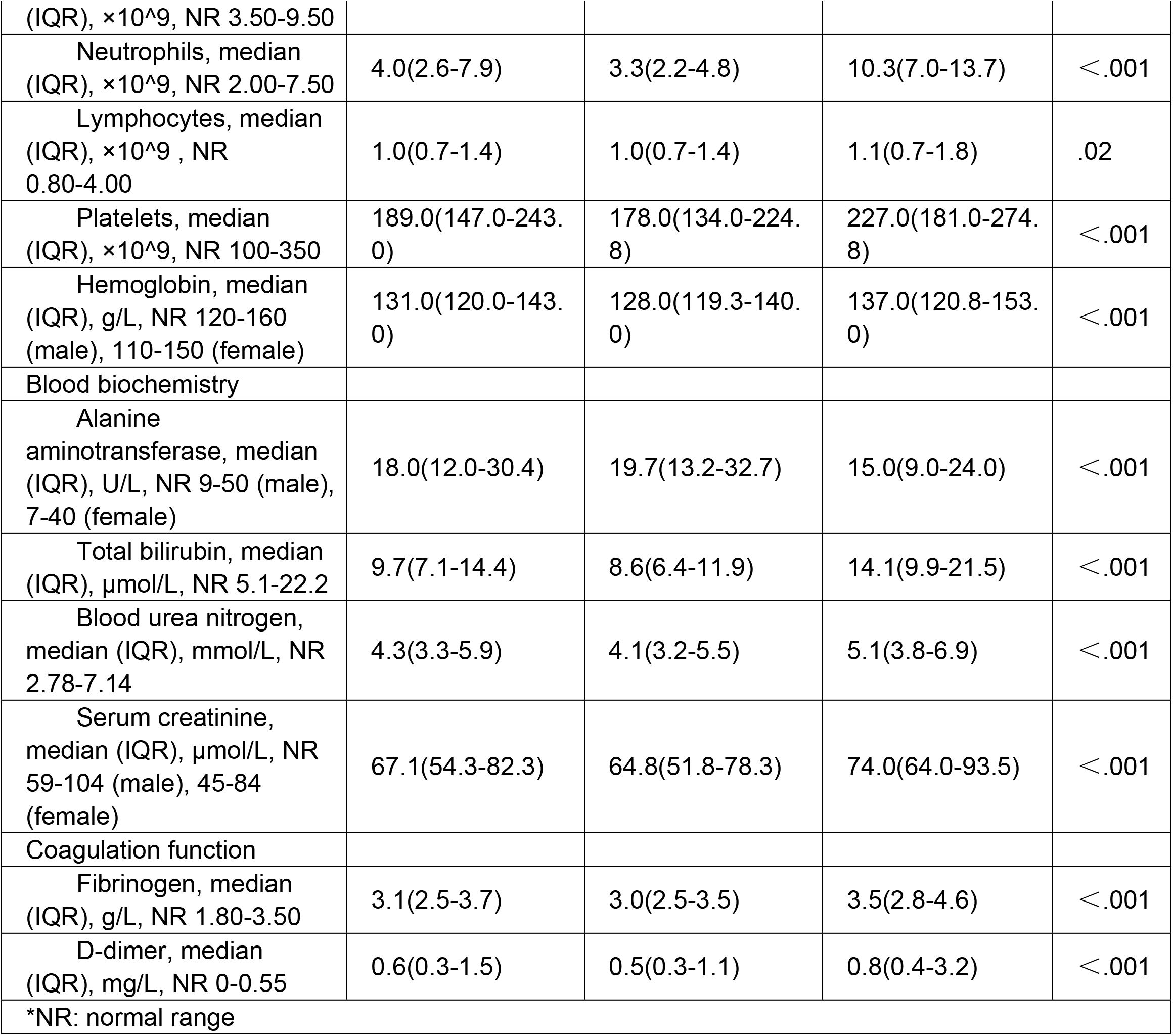
Demographic, Clinical, Imaging and Laboratory Characteristics of Patients on Admition or First to Emergency.

**Figure 1.**
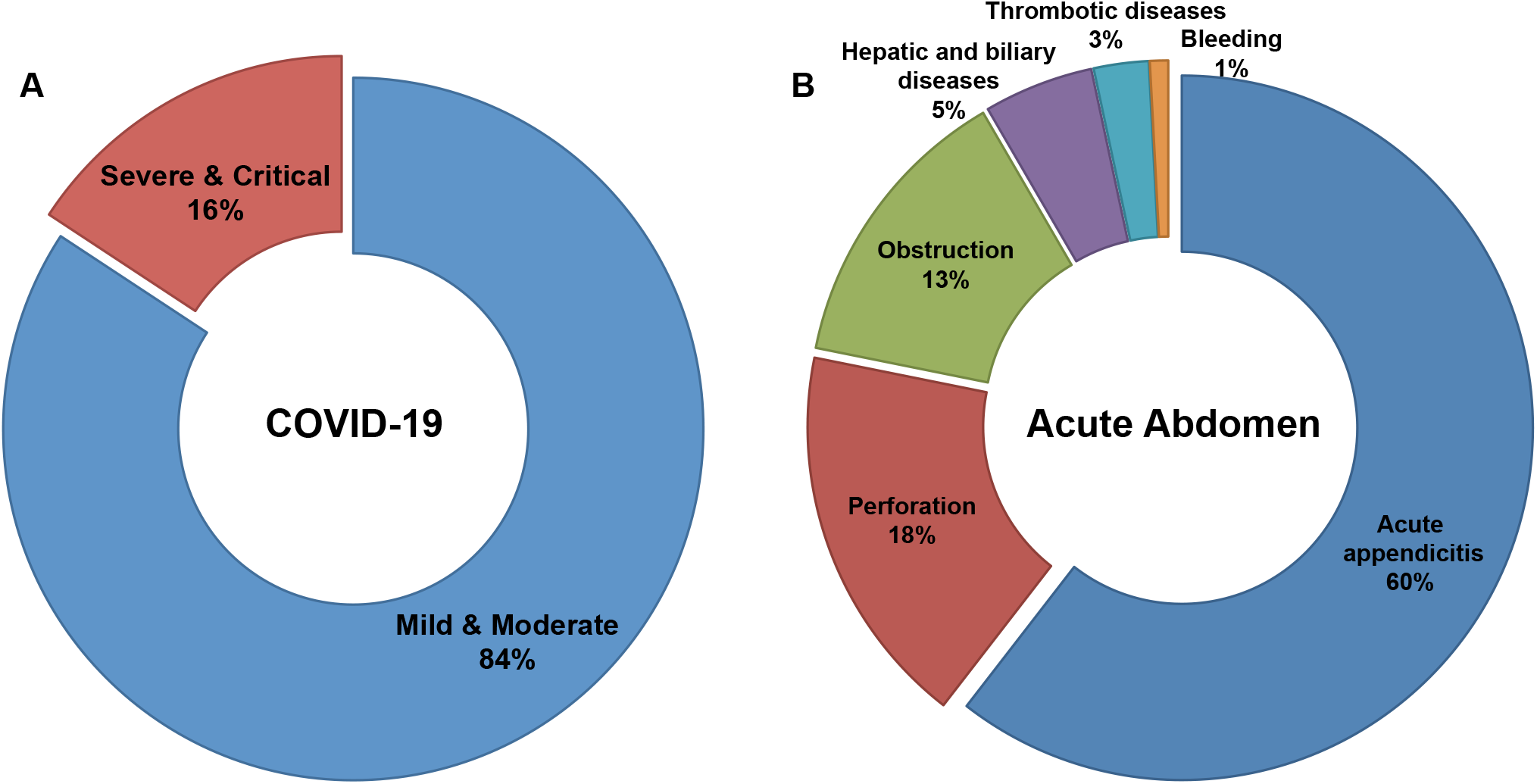
Distribution of severity in COVID-19 patients and disease spectrum in infectious acute abdomen patients. (A). 16% of enrolled COVID-19 patients were severe or critical and the left 84% were mild or moderate. (B). The disease spectrum of enrolled infectious acute abdomen patients showed the top 3 causes for emergency operation were acute appendicitis (60%), gastrointestinal perforation (18%) and bowel obstruction (13%).

### Potential predictors selection

All collected 40 variables were reduced to 6 potential predictors (abdominal pain, fever, chest CT, CRP, PCT and WBC) with nonzero coefficients in LASSO regression on the basis of 547 patients in the training cohort (Figure 2). It should be noted that 99.2% of the infectious acute abdomen patients had the symptom of abdominal pain, the proportion of which in COVID-19 patients was merely 0.2%, making it very likely to have overmuch weight in the model and go against the distinguish efficacy (Table 1). Concerned above, abdominal pain was excluded from the potential predictors. AUC of the left five variables, fever, chest CT, CRP, PCT and WBC, was yielded respectively in the training cohort and validation cohort (Figure S1 and S2).

**Figure 2.**
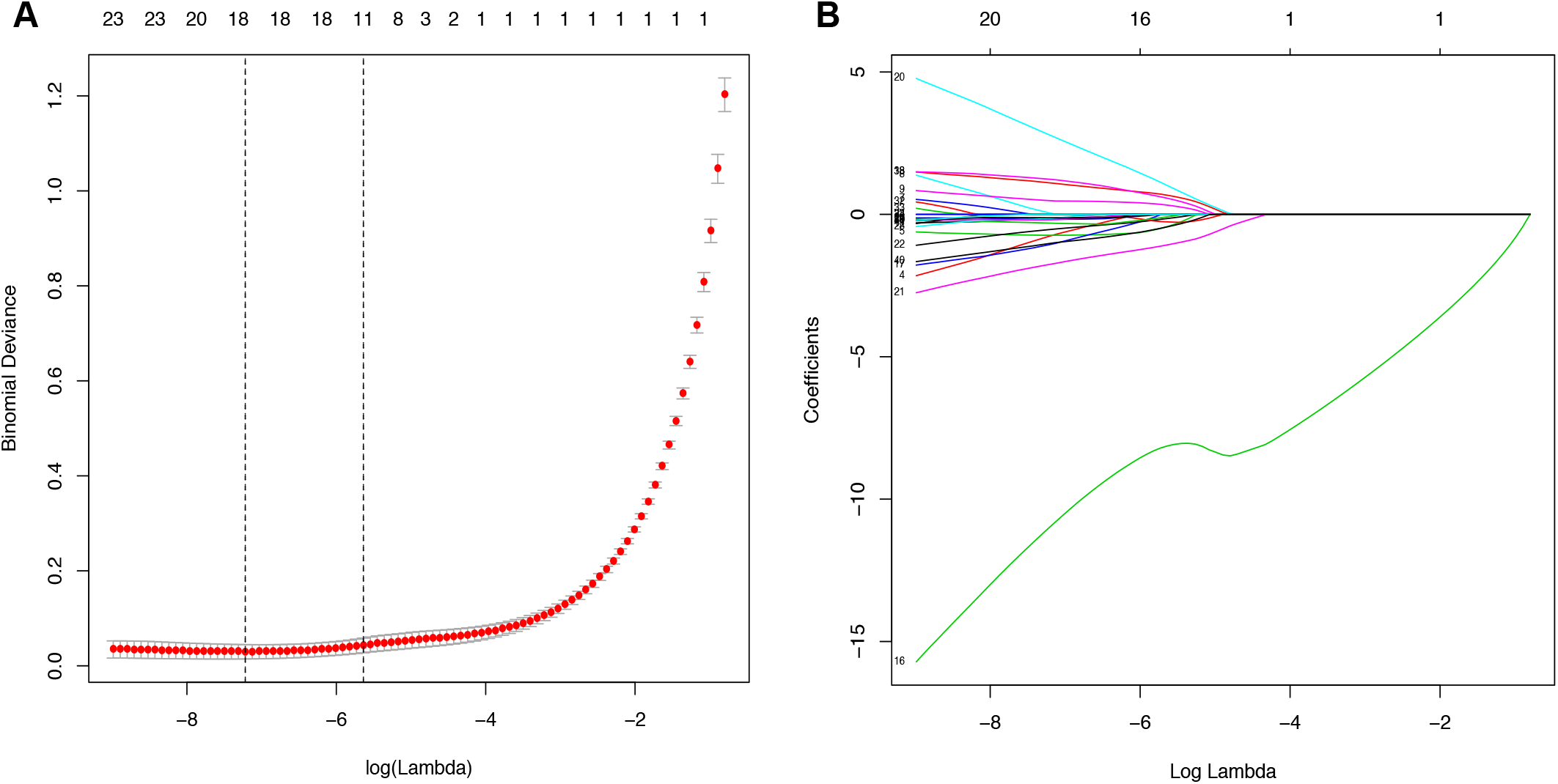
Potential predictors selection using the least absolute shrinkage and selection operator (LASSO) regression model. (A). The binomial deviance curve was plotted versus log(λ). Dotted vertical lines were drawn at the optimal values by using the minimum criteria and the 1 standard error of the minimum criteria (the 1-SE criteria). (B). LASSO coefficient profiles of the 40 alternative variables. A coefficient profile plot was produced against the log (λ) sequence.

### Development of a prediction model

To simplify the model, the concrete values of CRP, PCT and WBC were transformed into categorical variables (CRP and PCT: 1 for normal, 2 for high; WBC: 1 for low, 2 for normal and 3 for high) and the symptom fever was also defined as 0 for “no” and 1 for “yes”. Multivariate logistic regression analysis was performed among the five variables and all of these potential predictors have great value in distinguishing COVID-19 from infectious acute abdomen (Table 2). A risk score formula was preliminarily built to predict COVID-19 probability as follows: Logit (P = COVID-19) = 10.104 + 1.915×fever + 1.753×chest CT + (-2.508)×CRP + (-0.8)×PCT + (-1.836)×WBC. The CIAAD nomogram was generated on basis of the above result (Figure 3A).

**Table 2.**
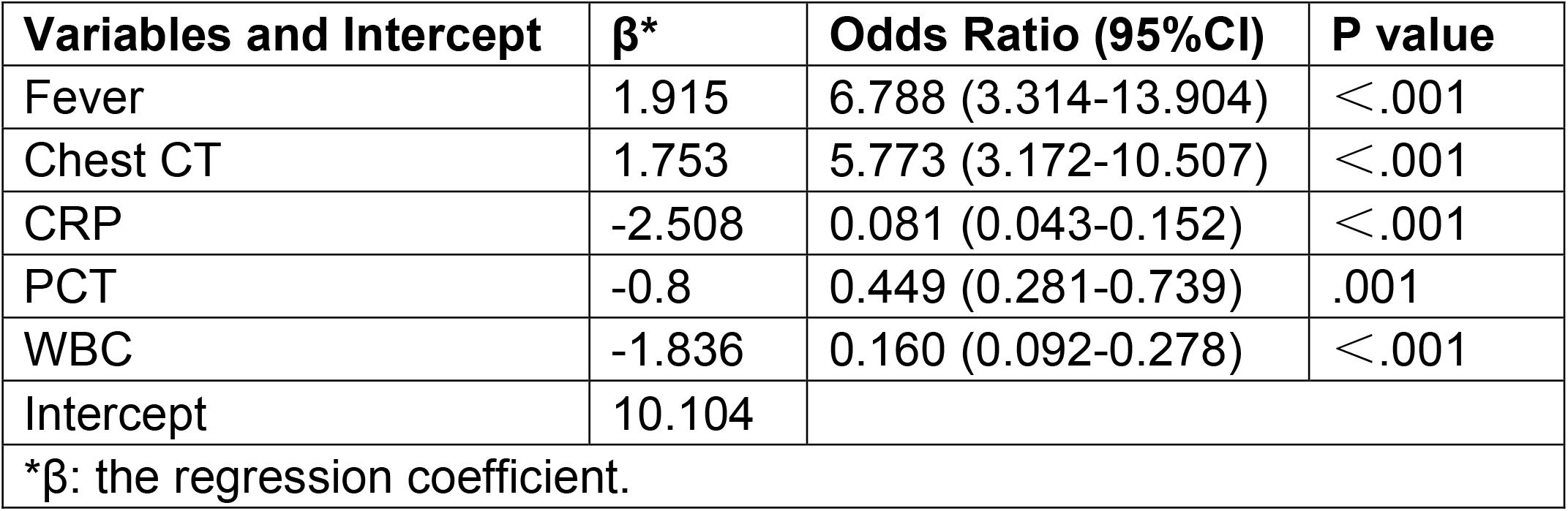
Multivariable Logistic Regression of Potential Predictors for screening COVID-19 in Infectious acute abdomen Patients (Training Cohort).

| **Variables and Intercept** | **$\beta^*$** | **Odds Ratio (95%CI)** | **P value** |
| --- | --- | --- | --- |
| Fever | 1.915 | 6.788 (3.314-13.904) | <.001 |
| Chest CT | 1.753 | 5.773 (3.172-10.507) | <.001 |
| CRP | -2.508 | 0.081 (0.043-0.152) | <.001 |
| PCT | -0.8 | 0.449 (0.281-0.739) | .001 |
| WBC | -1.836 | 0.160 (0.092-0.278) | <.001 |
| Intercept | 10.104 |  |  |
| *$\beta$: the regression coefficient. | | | |

**Figure 3.**
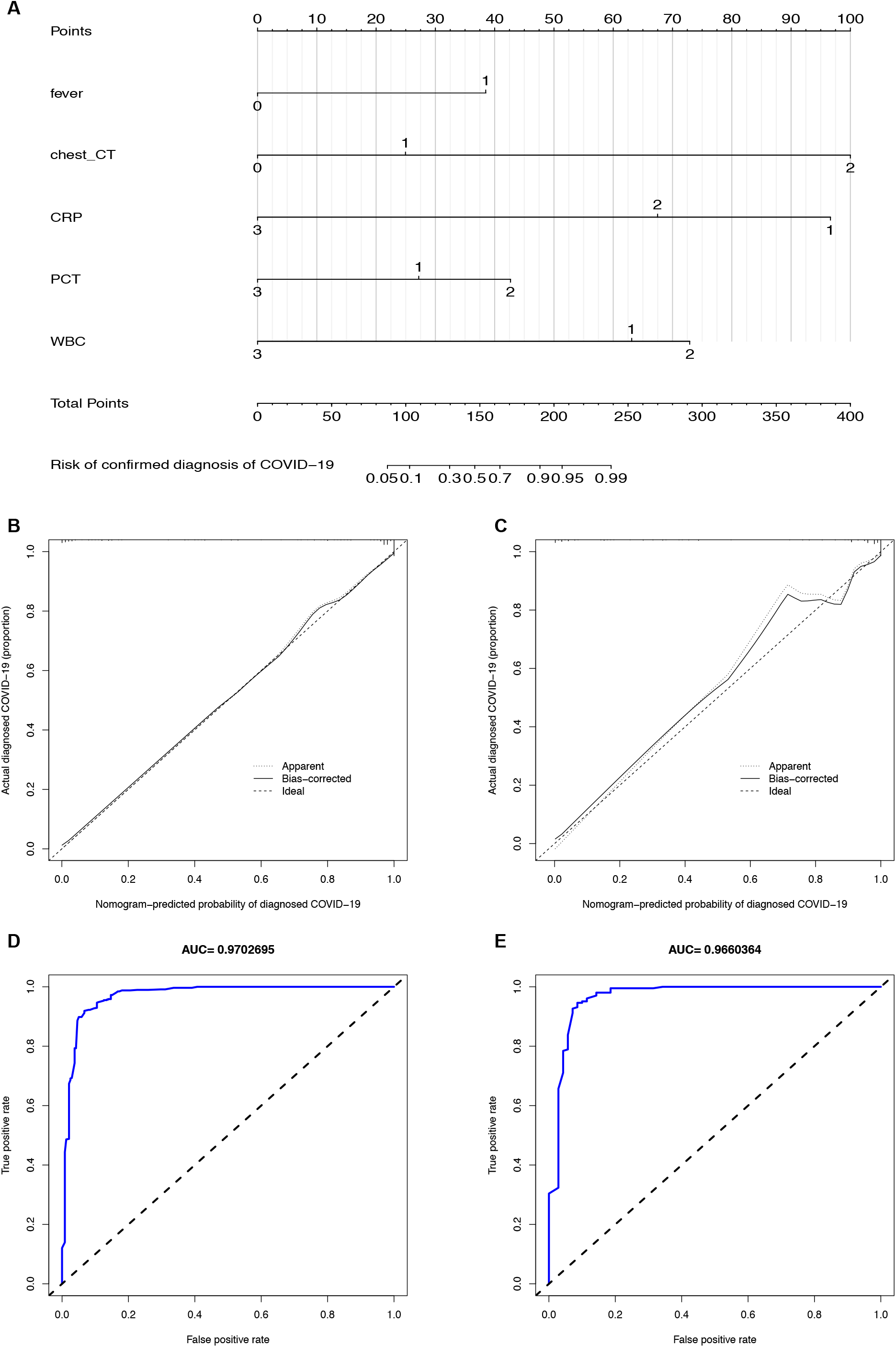
The CIAAD nomogram and its discrimination performance in training and validation cohort. (A). The CIAAD nomogram was developed in the training cohort based on fever, chest CT, CRP, PCT and WBC. (B). Calibration curve of CIAAD nomogram in the training cohort. Calibration curves depict the calibration of each model in terms of the agreement between the predicted risks of COVID-19 and observed outcomes of confirmed diagnosis of COVID-19. The Y-axis represents the actual COVID-19 rate. The X-axis represents the predicted COVID-19 risk. The diagonal dotted line represents a perfect prediction by an ideal model. The black solid line represents the performance of CIAAD nomogram, of which a closer fit to the diagonal dotted line represents a better prediction. (C). Calibration curve of CIAAD nomogram in the validation cohort. (D). ROC curve of CIAAD nomogram in the training cohort. (E). ROC curve of CIAAD nomogram in the validation cohort.

### Performance of the nomogram in training and validation cohort

The calibration curve of CIAAD nomogram for the risk of COVID-19 demonstrated good agreement between prediction and reality in the training cohort (Figure 3B). The C-index for the prediction nomogram was 0.981 (95% CI, 0.963 to 0.999) for the training cohort. Good calibration was also observed in the validation cohort with a C-index of 0.966 (95% CI, 0.960 to 0.972) (Figure 3C). The ROC analysis for the training and validation cohort yielded AUC values of 0.970 (95% CI, 0.961 to 0.982) and 0.966 (95% CI, 0.957 to 0.975), which implied the prediction performance was favorable (Figure 3D and 2E).

### Clinical Use and development of a simplified scale

Decision curve analysis and clinical impact curves was conducted for the CIAAD nomogram in both training and validation cohort (Figure 4), demonstrating high clinical net benefit nearly over the entire threshold probability.

**Figure 4.**
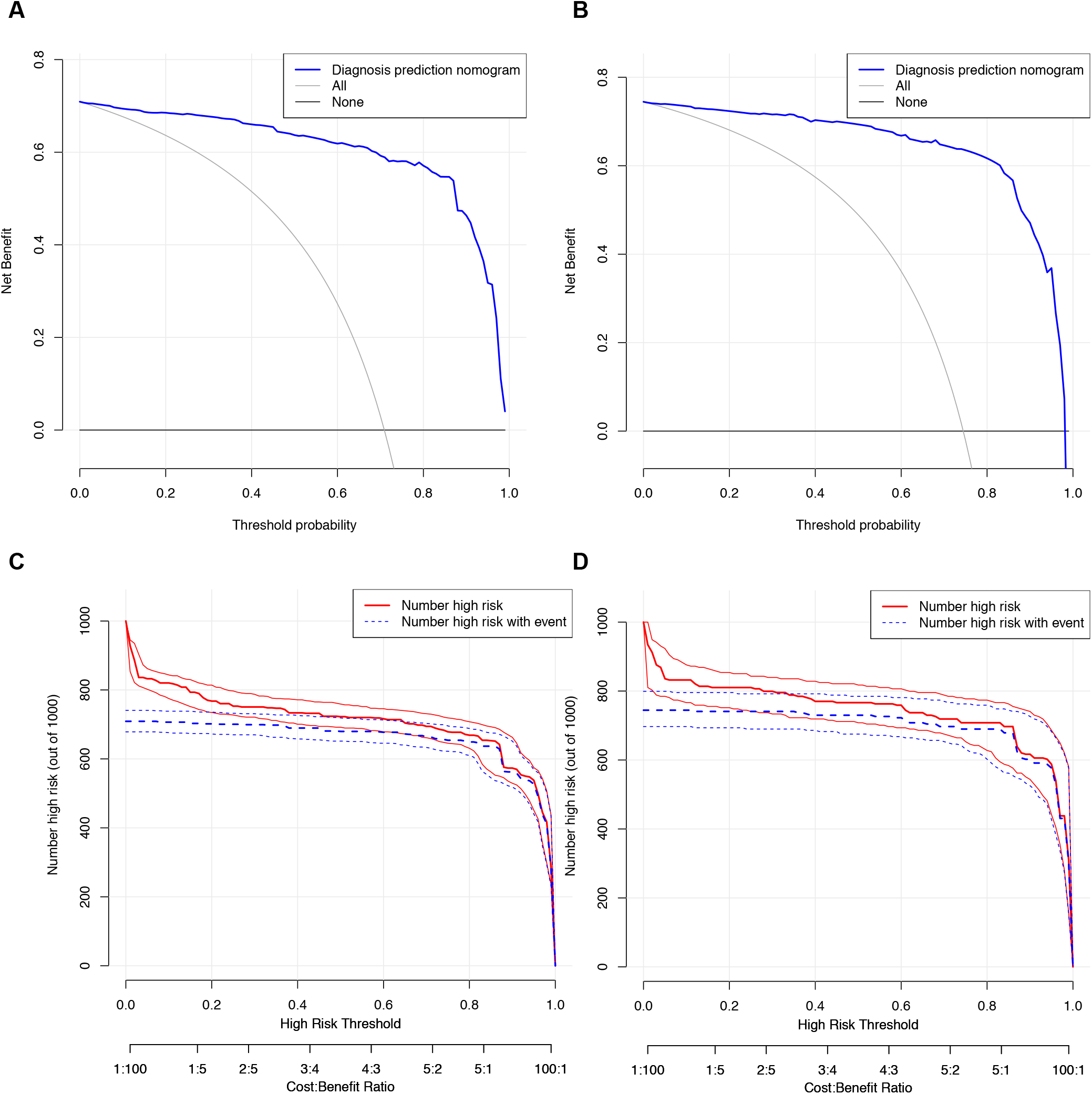
Decision curve analysis and clinical impact curves for CIAAD nomogram in training and validation cohort. (A). Decision curve analysis for CIAAD nomogram in the training cohort. The Y-axis measures the net benefit. The blue line represents the CIAAD nomogram. The grey line represents the assumption that all patients are COVID-19 patients. The black line represents the assumption that there are no COVID-19 patients. (B). Decision curve analysis for CIAAD nomogram in the validation cohort. (C). Clinical impact curve for CIAAD nomogram in the training cohort. The red curve (Number high risk) indicates the number of people classified as positive (high risk) by nomogram under each threshold probability. The blue curve (Number high risk with event) is the number of truly positive people under each threshold probability. (D). Clinical impact curve for CIAAD nomogram in the validation cohort.

With the purpose of making our prediction model more concise and practical in surgery emergency, we simplified the scoring criterion of the CIAAD nomogram and create a brand new scale, named CIAAD scale (Figure 5). In CIAAD scale, the lowest and highest score of this scale is 3.5 and 14.5. The item with a higher corresponding score is more common in COVID-19 patients, such as fever, abnormal chest CT, normal level of CRP and WBC. If the total score of a patient is less than 5, he/she gets low risk (<30%) of confirmed COVID-19, and the risk rises to more than 70% as the total score reaches 7.

**Figure 5.**
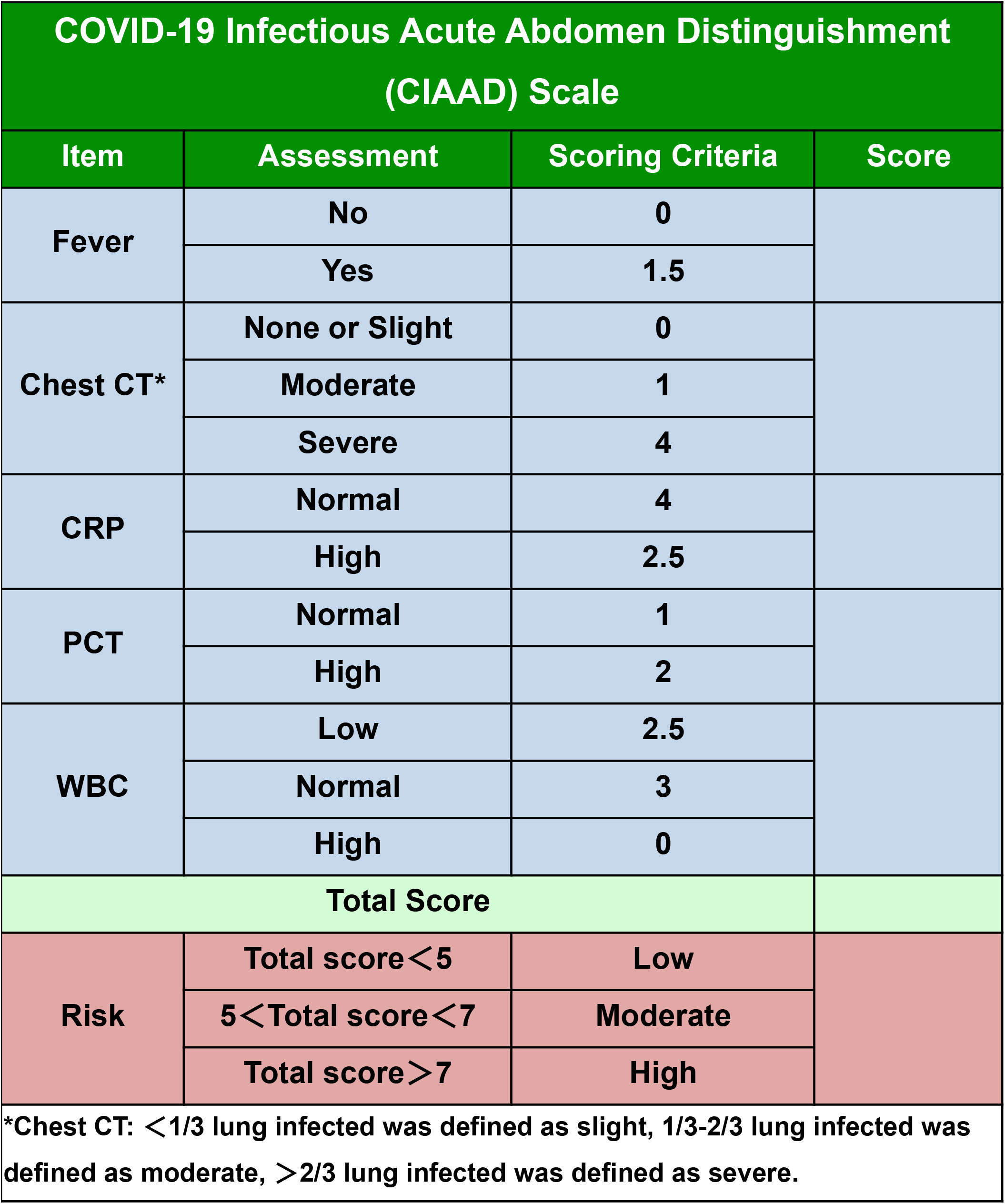
The COVID-19 Infectious Acute Abdomen Distinguishment Scale based on CIAAD nomogram. Patients with a total score less than5 were considered of low risk of true SARS-CoV-2 infection, 5 to 7 of moderate risk and more than 7 of high risk.

## Discussion

With the global outbreak of COVID-19, the latest sum of infected patients has exceeded 1.8 million, and humankind will face the threat all through the near future^2^. Vast medical resource has been put into rolling back the virus resulting in the treatment for many other diseases postponed. However, for surgeons confronted with patients of infectious acute abdomen urgent for emergency operation, it is necessary to accurately distinguish COVID-19 patients from the ones with similar and misguiding symptoms in the shortest time. To prevent cross-infection of medical staff, doctors, nurses and other patients in outpatient clinic, wards and operating rooms should take high-level precaution when managing COVID-19 highly suspected patients. Excessive precaution would cause huge waste of precious medical resource. On the contrary, negligence of necessary screening would fling medical staff into great risk of infection. The current screening procedures, such as nucleicacid detection and CT, have the defects of unsatisfactory accuracy and time-consuming. Consequently, a more convenient, efficient, economical and effective COVID-19 screening method is surgeons’ desideratum. To our knowledge, this study provided the first screening model and scale for surgeons to distinguish infectious acute abdomen patients from suspected and high risk of COVID-19 patients in emergency department by retrospectively comparing demographic, clinical and laboratory characteristics of the two groups of patients. The CIAAD nomogram and scale has satisfying performance in prediction and great potential to help medical institutions to resume routine medical work during the epidemic.

### Challenges and opportunities

Current reports of the COVID-19 cohorts show that respiratory symptoms, such as fever, cough and dyspnea, are the main clinical manifestations^10^. Nevertheless, the digestive symptoms like diarrhea, nausea, vomiting and abdominal pain are gradually reported to be an early onset, which deserves more attention^11,12^. A meta-analysis revealed that nearly half of the patients had positive results for SARS-CoV-2 in stool and another bioinformatics analysis had provided probable theoretical basis for the digestive symptoms, that angiotensin converting enzyme II (ACE2) was highly expressed in esophagus, ileum and colon^13,14^. The mixture of fever and some digestive symptoms mimics the symptoms of infectious acute abdomen to a great extent. Similarly, if the patient is elder or the abdominal infection develops into systemic infection, signs of pneumonia will emerge in infectious acute abdomen patients as well. For infectious acute abdomen, increased morbidity and mortality associated with a delay in the treatment of many of the surgical causes argue for an aggressive and expeditious surgical approach^15^. In the epidemic, quick and accurate screening in infectious acute abdomen patients suspected of COVID-19 is vitally important.

The definite diagnosis of COVID-19 is still mainly depends on a positive RT-PCR result for SARS-CoV-2^16^, which has the shortage of possibility of false-negative caused by various reasons, such as disease stage, virus load and sample quality. In our COVID-19 cohort, the positive rate at the first time of nucleicacid detection is merely 43.7% (255/584). Meanwhile, chest CT was also considered as a good tool to screen. However, mild patients without pneumonia, atypical imaging findings and great dependence on physicians’ experience limit the screening value of chest CT. A study enrolled in 1014 COVID-19 cases from Wuhan showed that the positive rate for chest CT was 88%, comparing the 59% of RT-PCR^8^. Several prediction models based on the integration of demographic, clinical, imaging and laboratory variables have been developed to evaluate the disease risk or prognosis^17^. Unfortunately, the target population of published diagnostic models was patients presenting at fever clinic or ordinary patients suspected COVID-19^17^. It is not suitable for surgeons to borrow these models directly to screen infectious acute abdomen patients and our CIAAD nomogram and scale make up the pity.

### Strengths and limitations of this study

The set of our model has the superiority of strong pertinence. The recommended user of CIAAD scale is surgeon in the emergency department and the recommended assessed population is infectious acute abdomen patients suspected COVID-19. To this end, we collected firsthand and high-quality data of COVID-19 patients and enrolled infectious acute abdomen patients strictly. In addition, by virtue of LASSO regression analysis, five quantifiable indicators were successfully selected. Though many variables like diabetes, cough and D-dimer varied considerably between COVID-19 and infectious acute abdomen patients, they were ruled out by LASSO regression analysis as overmuch weight or causing the prediction model cumbersome. The selected indictors were all included in previous prediction models, which verified the prediction capacity of these variables from the side^17^. Whereas, there is some inadequacy in our study as well. Firstly, the disturbance for routine medical work by the epidemic resulted in the appropriate lack of patients with both COVID-19 and acute abdomen. As the number of emergency operations of acute abdomen decreased sharply in Wuhan, the data of acute abdomen patients was from Peking Union Medical College Hospital, a renowned and leader hospital in China.

### Implications for practice and future

An algorithm helpful for allowing both a focused workup and expeditious therapy was given, including necessary prevention advice for medical staff with the guidelines published by World Health Organization for reference^18^ (Figure 6). What needs to be pointed out is that standard precautions are needed for all patients. For an infectious acute abdomen patient suspected COVID-19, the first step is to evaluate his/her surgical status and screen the patient by CIAAD scale. The urgency degree of surgical status determines whether the medical staff wait for the results of nucleicacid detection or take precautions according to our algorithm. Level of precautions adopted should be instructed by the risk degree of COVID-19 from CIAAD scale. As the scale is harmless and the has net benefit nearly over the entire threshold probability according to the decision analysis curves, strong recommendation of our CIAAD scale and the algorithm was made to all surgeons in countries severely affected by the epidemic. With the wide use in lager population, efficacy of CIAAD scale will be further tested in a prospective way.

**Figure 6.**
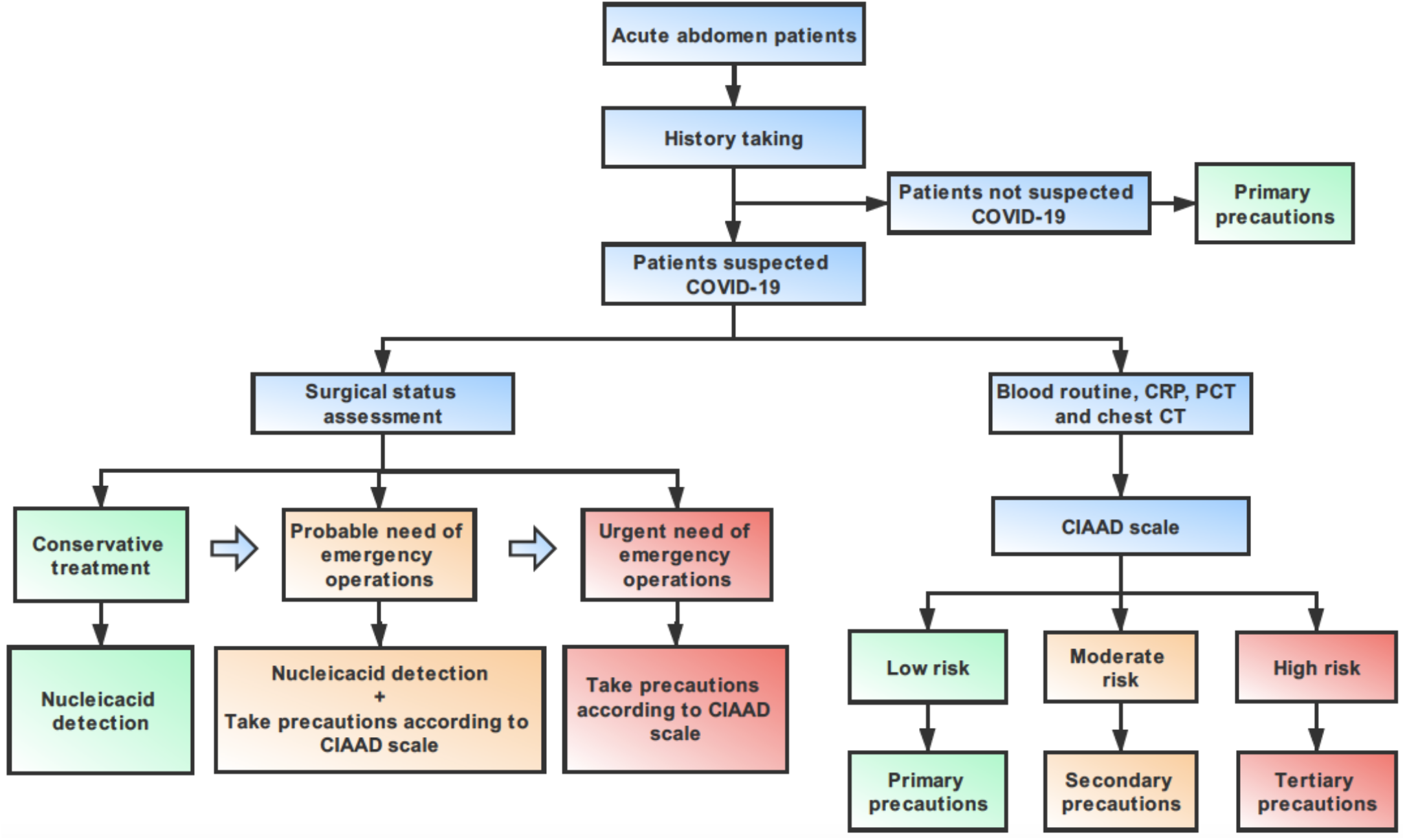
An algorithm help surgeons to manage infectious acute abdomen patients suspected COVID-19 in the emergency department (ED). Key procedures in this algorithm are history taking, surgical status assessment and CIAAD scale. Note: Primary precautions are needed for all patients in ED during the epidemic.

## Conclusion

With the aim of distinguishing COVID-19 patients from infectious acute abdomen patients, we established an easy and effective screening model and scale for surgeons in emergency department. The algorithm based on CIAAD scale is promising to help surgeons manage infectious acute abdomen patients suspected COVID-19 more efficiently avoiding cross infection as well as guard health of medical staff.

## Data Availability

There is no supplementary data available referred to in the manuscript.

## ARTICLE INFORMATION

### Author Contirbutions

ZBB, WYX, SWW and QC contributed equally to this article. WWB, WYJ, ZBB, WYX and SWW designed the study. ZBB, WYX, SWW, ZXT, LTH, CHT and WWB collected assembled the data. ZBB, WYX, SWW, QC and WZH analysed and interpreted the data. ZBB, WYX and SWW wrote the first draft, which all authors revised for critical content. All authors approved the final manuscript. WWB and WYJ are the guarantors. The corresponding authors attest that all listed authors meet authorship criteria and that no others meeting the criteria have been omitted.

### Ethical approval

This study was approved by the Ethics Committees of the Peking Union Medical College Hospital and the Central Hospital of Wuhan.

### Patient consent

Not applicable.

### Transparency

The lead authors and manuscript’s guarantor affirm that the manuscript is an honest, accurate, and transparent account of the study being reported; that no important aspects of the study have been omitted; and that any discrepancies from the study as planned have been explained.

### Dissemination to participants and related patient and public communities

No patients or members of the public involved in the design, or conduct, or reporting, or dissemination plans of this study.

---

**Supplementary figure 1.**
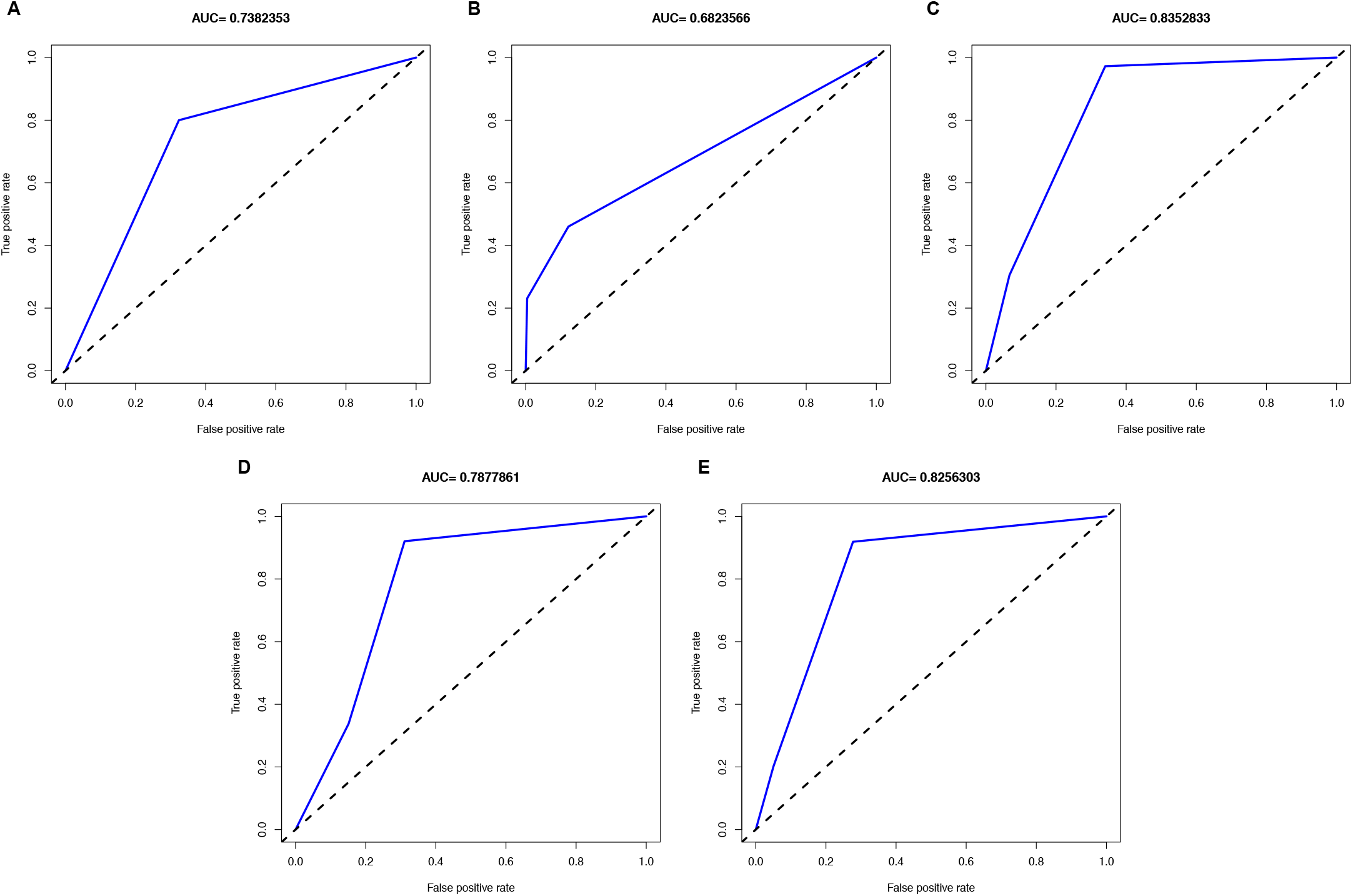
ROC curves of each potential predictors in training cohort. (A) - (E) for fever, chest CT, CPR, PCT and WBC respectively.

**Supplementary figure 2.**
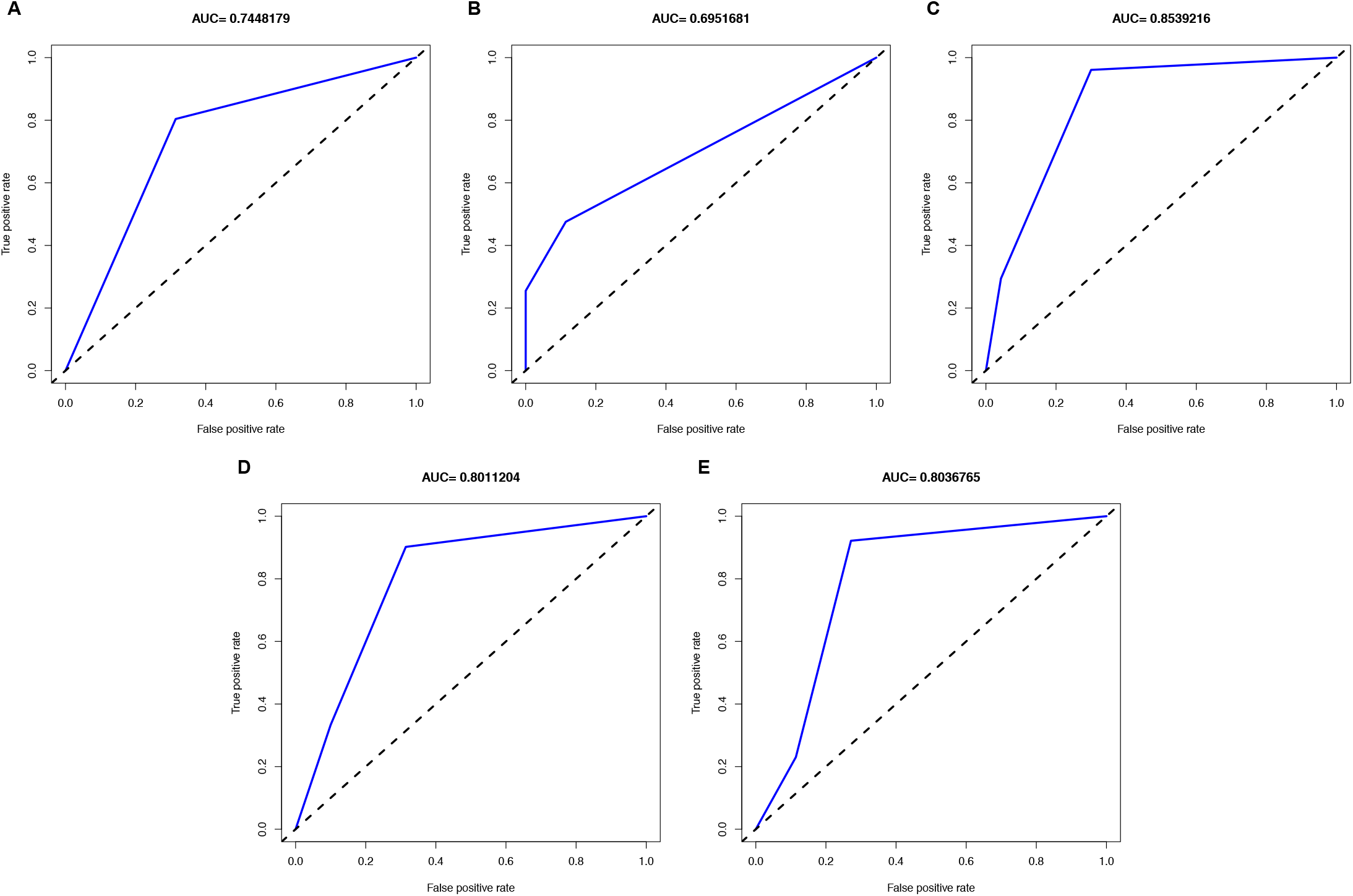
ROC curves of each potential predictors in validation cohort. (A) - (E) for fever, chest CT, CPR, PCT and WBC respectively.

